# Longitudinal cellular and humoral immune responses after triple BNT162b2 and fourth full-dose mRNA-1273 vaccination in haemodialysis patients

**DOI:** 10.1101/2022.07.13.22277581

**Authors:** Matthias Becker, Anne Cossmann, Karsten Lürken, Daniel Junker, Jens Gruber, Jennifer Juengling, Gema Morillas Ramos, Andrea Beigel, Eike Wrenger, Gerhard Lonnemann, Metodi V. Stankov, Alexandra Dopfer-Jablonka, Philipp D. Kaiser, Bjoern Traenkle, Ulrich Rothbauer, Gérard Krause, Nicole Schneiderhan-Marra, Monika Strengert, Alex Dulovic, Georg M.N. Behrens

**Affiliations:** NMI Natural and Medical Sciences Institute at the University of Tübingen, Reutlingen, Germany; Department for Rheumatology and Immunology, Hannover Medical School, Hannover, Germany; Dialysis Centre Eickenhof, Langenhagen, Germany; CiiM - Centre for Individualized Infection Medicine, Hannover, Germany; Pharmaceutical Biotechnology, University of Tübingen, Germany; German Centre for Infection Research (DZIF), partner site Hannover-Braunschweig, Germany; Helmholtz Centre for Infection Research, Braunschweig, Germany; TWINCORE GmbH, Centre for Experimental and Clinical Infection Research, a joint venture of the Hannover Medical School and the Helmholtz Centre for Infection Research, Hannover, Germany

**Keywords:** SARS-CoV-2, dialysis, mRNA vaccination, Omicron variant of concern, protective immunity, immunocompromised, longitudinal response, heterologous immunisation

## Abstract

**Background:** Haemodialysis patients are at-risk for severe COVID-19 and were among the first to receive a fourth COVID-19 vaccination.

**Methods:** We analysed humoral responses by multiplex-based IgG measurements against the receptor-binding domain (RBD) and ACE2-binding inhibition towards variants of concern including Omicron in haemodialysis patients and controls after triple BNT162b2 vaccination and in dialysis patients after a fourth full-dose of mRNA-1273. T-cell responses were assessed by interferon γ release assay.

**Findings:** After triple BNT162b2 vaccination, anti-RBD B.1 IgG and ACE2 binding inhibition reached peak levels in dialysis patients, but remained inferior compared to controls. Whilst we detected B.1-specific ACE2 binding inhibition in 84% of dialysis patients after three BNT162b2 doses, binding inhibition towards the Omicron variant was only 38% and declining to 16% before the fourth vaccination. By using mRNA-1273 as fourth dose, humoral immunity against all SARS-CoV-2 variants tested was strongly augmented with 80% of dialysis patients having Omicron-specific ACE2 binding inhibition. Modest declines in T-cell responses in dialysis patients and controls after the second vaccination were restored by the third BNT162b2 dose and significantly increased by the fourth vaccination.

**Conclusions:** A fourth full-dose mRNA-1273 after triple BNT162b2 vaccination in haemodialysis patients leads to efficient humoral responses against Omicron. Our data support current national recommendation and suggest that other immune-impaired individuals may benefit from this mixed mRNA vaccination regimen.

**Funding:** Initiative and Networking Fund of the Helmholtz Association of German Research Centres, EU Horizon 2020 research and innovation program, State Ministry of Baden-Württemberg for Economic Affairs, Labour and Tourism, European Regional Development Fund

**Research in the context:** *Evidence before this study:* Information on how to best maintain immune protection after SARS-CoV-2 vaccination in at-risk individuals for severe COVID-19 such as haemodialysis patients is limited. We searched PubMed and medRxiv for keywords such as “haemodialysis”, “SARS-CoV-2”, “vaccine”, “decay”, “antibody kinetics”, “cellular immunity”, “longitudinal vaccination response”, “immunisation scheme”. To date, no peer-reviewed studies comprehensively assessed impact of both cellular and humoral immunogenicity after a triple BNT162b2 vaccination in combination with a fourth full-dose of mRNA-1273 and addressed the impact of currently dominating SARS-CoV-2 variants of concern on vaccine-induced immunity in this at-risk population.

*Added value of the study:* We provide to the best of our knowledge for the first time longitudinal vaccination response data over the course of the pandemic in dialysis patients. We studied not only systemic T- and B-cell but also mucosal responses in this at-risk group and determined levels of neutralizing antibodies towards Omicron BA.1 and Delta variants after a mixed mRNA vaccine schedule.

*Implications of all the available evidence:* Patients on haemodialysis show inferior response rates and thus a more rapid decline in humoral immune response after triple vaccination with BNT162b2. Our data strongly support the concept of administering a fourth full-dose of mRNA-1273 as part of a heterologous vaccination scheme to boost immunity and to prevent severe COVID-19 within this at-risk population. Strategic application of modified vaccine regimens may be an immediate response against SARS-CoV-2 variants with increased immune evasion potential.

## 1. Introduction

To date, SARS-CoV-2 vaccinations reassuringly provide some degree of protection from severe COVID-19 independent of the currently circulating variants of concern (VoC) for the majority of healthy individuals (1). However, weaker immunogenicity and a faster decline in protection levels to standard two-dose or three-dose booster SARS-CoV-2 immunisation schemes have been widely demonstrated in immunocompromised individuals such as solid organ transplant recipients (2), dialysis patients (3) or patients suffering from other severe chronic conditions such as cancer (4). Already in mid-2021 or then in the beginning of 2022, several countries recommended a fourth dose of SARS-CoV-2 mRNA vaccines not only for immunosuppressed populations at risk for severe COVID-19 disease, but also for older individuals to increase and maintain levels of immune protection (5-8). This was driven by both weaker peak vaccine responses and waning immunity in this individuals as well as continued evolution of SARS CoV-2 variants with increasing levels of immune evasion potential as demonstrated by Omicron VoC subspecies BA.1, BA.4, BA.5, and BA.2.12.1 (9-12).

Recent studies reported an improved SARS-CoV-2 humoral and cellular responses not only towards the original SARS-CoV-2 B.1 isolate but also Delta and Omicron VoC after a fourth vaccination in haemodialysis patients receiving either mRNA vaccines or vector-based formulations in combination with mRNA vaccines (13-15). However, targeted data on the most efficient dosing and vaccination scheme or even predictors of vaccination success in haemodialysis patients at-risk of severe COVID-19 and its associated mortality is limited. We aimed to comprehensively examine the magnitude and kinetics of both cellular and humoral immunity towards the most recently dominating Delta and Omicron variant’s in a well-controlled longitudinal cohort of haemodialysis patients. These patients received a triple dose of BNT162b2 followed by a fourth full-dose of mRNA-1273. Healthcare workers vaccinated three time with BNT162b2 served as controls. Our data provide preliminary evidence that in addition to heterologous vector- and mRNA-based vaccination schemes also heterologous mRNA vaccine regimens maybe strategically beneficial for achieving efficient immunity against SARS-CoV-2 in immunosuppressed patients.

## 2. Methods

### 2.1 Study design and sample collection

This is a follow-up study in haemodialysis patients and control individuals, for which the results for haemodialysis patients after a complete two-dose BNT162b2 vaccination (16) and subsequent decline (17) have been previously reported. Blood samples were taken before start of dialysis (n=50) or from healthcare workers (n=33), who participated in the COVID-19 contact (CoCo) study served (18) as non-dialysed control population. To be included in the study, participants had to be over the age of 18 and able to give written informed consent. For the current analysis, we only considered dialysis patients for which results from all time points after either three or four vaccine doses were available. All participants received the standard two-dose regimen of BNT162b2 three weeks apart, followed by a third BNT162b2 vaccination about six (dialysis) or 8·5 months (controls) after the second vaccination. Only dialysis patients were vaccinated a fourth time with 100 µg mRNA-1273 four months after the last BNT162b2 vaccination. The vaccination schedule and blood collection time points are depicted in Fig. 1 and Fig. S1. Participants with SARS-CoV-2 infection diagnosed by either PCR or anti-nucleocapsid IgG determined by MULTICOV-AB multiplex measurement (19) were excluded from the analysis. Demographic characteristics and medical information are listed in Table 1, S1 and S2. Plasma was obtained from lithium heparin blood (S-Monovette Plasma, Sarstedt, Germany). Whole blood samples were used immediately for interferon γ release assay (IGRA). For saliva collection, all individuals spat directly into a collection tube. To inactivate replication-competent SARS-CoV-2 virus particles potentially present in saliva samples, Tri(n-butyl) phosphate (TnBP) and Triton X-100 were added to final concentrations of 0·3% and 1%, respectively (20). Both plasma and saliva samples were frozen and stored at −80°C until further use.

**Table 1.** Characteristics of study population. IQR - Inter Quartile Range. BMI - Body Mass Index. n – absolute numbers per group. NA - Information not available. n. a. - not applicable.

| Characteristics | Haemodialysis group (n=50) | Non-dialysis control group (n=33) | p-value for difference between groups |
| --- | --- | --- | --- |
| Age (years), median (IQR) | 69·5 (60-79) | 42 (32-55) | 1·08*10<sup>-11</sup> |
| Sex (female: n, %) | 19 (38·0) | 23 (69·7) | 9·26*10<sup>-3</sup> |
| Days since start of haemodialysis (median, IQR) | 1263 (753-2314) | n. a. | n. a. |
| Immunosuppressive medication (n, %) |  |  |  |
| 2021 (Vaccine dose 1-3)* | 7 (14·0) | 0 (0·0) | n. a. |
| 2022 (Vaccine dose 4)* | 6 (12·0) | n. a. | n. a. |
| Co-morbidities |  |  |  |
| Obesity (BMI, >30) | 12 (24·0) | NA | n. a. |
| Diabetes mellitus (n, %) | 14 (28·0) | 1 (3·0) | 9·27*10<sup>-3</sup> |
| Cardiovascular disease (n, %) | 21 (42·0) | 2 (6·1) | 8·69*10<sup>-4</sup> |
| Cancer (n, %) | 1 (2·0) | 0 (0·0) | n. a. |
| Chronic conditions (n, %) |  |  |  |
| Ulcerative colitis (n, %) | 0 (0·0) | 1 (3·0) | n. a. |
| Goiter (n, %) | 0 (0·0) | 1 (3·0) | n. a. |
| Hashimoto's thyroiditis (n, %) | 0 (0·0) | 1 (3·0) | n. a. |
| Hypothyroidism (n, %) | 0 (0·0) | 1 (3·0) | n. a. |
| Other | 0 (0·0) | 1 (3·0) | n. a. |

\* on medication when vaccinated and sampled

**Fig. 1.**
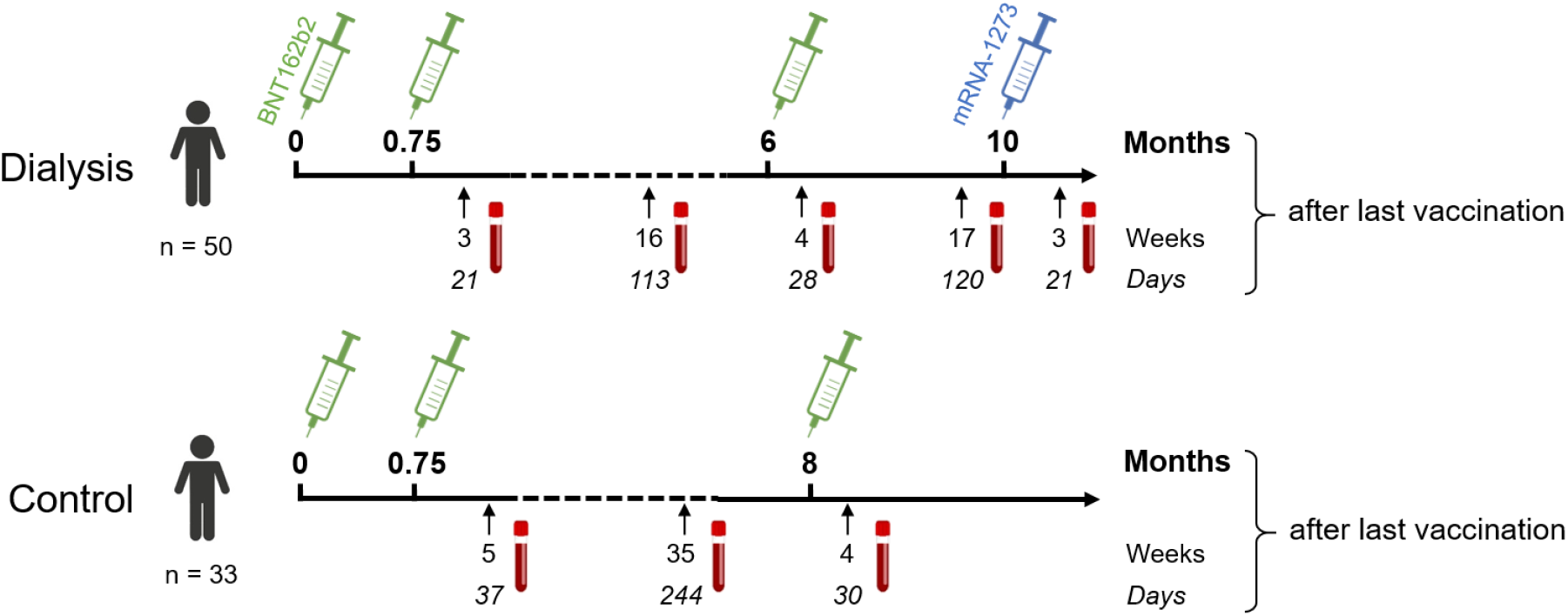
Participant recruitment scheme for longitudinal vaccination response analysis in haemodialysis patients after triple BNT162b2 and fourth full-dose mRNA-1273. Patients on haemodialysis (n=50) and healthcare workers as controls (n=33) were triple-vaccinated with BNT162b2 (green syringe) followed by a 100 µg (full) dose of mRNA-1273 (blue syringe) for dialysed individuals only. Sampling and vaccination schedule is given in days and weeks as indicated.

### 2.2 Ethics statement

Ethical approval was obtained from the Internal Review Board of Hannover Medical School (MHH, approval number 8973_BO-K_2020, amendment Dec. 2020). Written informed consent was obtained from all participants prior to starting the study.

### 2.3 MULTICOV-AB

IgG binding and levels were analysed using MULTICOV-AB, a multiplex coronavirus immunoassay which contains the trimeric Spike B.1, its subdomains (S1, S2, RBD), nucleocapsid B.1 and RBDs of Delta and Omicron BA.1 antigens as previously described (9, 19). Briefly, antigens were immobilised on spectrally distinct populations of MagPlex beads (Cat #MC10XXX-01, Luminex Corporation) either by EDC/s-NHS coupling (21) or by Anteo coupling (Cat #A-LMPAKMM-10, Anteo Tech Reagents) following the manufacturer’s instruction (19). The combined MagPlex beads were then incubated with samples at an effective dilution of 1:3200 for plasma and of 1:12 for saliva. After a wash step to remove unbound antibodies, IgG was detected with R-phycoerythrin labelled goat-anti-human IgG (Jackson ImmunoResearch Labs, Cat #109-116-098, Lot #148837, RRID: AB_2337678) as secondary antibody. After another wash step and bead resuspension, samples were measured once on a FLEXMAP 3D instrument (Luminex Corporation) using the following settings: Timeout 80 sec, Gate: 7500-15000, Reporter Gain: Standard PMT, 50 events. Raw median fluorescence intensity (MFI) values or normalised values (MFI/MFI of quality control (QC) samples (19, 22) are reported. Three QC samples were measured per individual plate to monitor MULTICOV-AB performance.

#### 2.4 RBDCoV-ACE2

RBDCoV-ACE2, a multiplex competitive inhibition assay, was performed as previously described (23) as surrogate assay to determine immunoglobulin neutralisation capacity against SARS-CoV-2 B.1 isolate and variants of concern. For this, biotinylated ACE2 was combined with individual samples (and as a control, ACE2 alone) and incubated with the above mentioned MULTICOV-AB bead mix. Before and after ACE2 detection with Strep-PE (Cat #SAPE-001, Moss), washes were carried out. Samples were measured once on a FLEXMAP 3D instrument with the same settings as MULTICOV-AB and analysed by normalisation of MFI values against the control. 100% ACE2 binding inhibition indicates maximum binding inhibition. Responders for ACE2 binding inhibition are classified as above a 20% ACE2 binding threshold as described in Junker *et al*. (23).

### 2.5 Anti-SARS-CoV-2 QuantiVac ELISA

Plasma samples were additionally analysed using the Anti-SARS-CoV-2-QuantiVac-ELISA IgG (Cat #EI 2606-9601-10G, Euroimmun) as previously described (16).

### 2.6 Interferonγ release assay

SARS-CoV-2-specific T-cell responses from whole blood were analysed by measuring IFNγ production after stimulation with a peptide pool from the SARS-CoV-2 Spike S1 with the SARS-CoV-2 Interferon Gamma Release Assay (Cat #ET-2606-3003, Euroimmun) and the IFNγ ELISA (Cat #EQ-6841-9601, Euroimmun) as previously described (16). Background signals from negative controls were subtracted and final results calculated in mIU/mL using standard curves. IFNγ concentrations >200 mIU/mL were considered as reactive. Results from positive and negative controls were not statistically significant different between time point T1 and T2. We defined this arbitrary cut-off by using average background IFNγ activity without antigen-stimulation in all samples of T1 multiplied with 10 for the threshold for IGRA-positive. Using this cut-off, we found in all of the 15 controls taken from independent individuals before the COVID-19 pandemic negative IGRA results (24). The upper limit of reactivity was 2000 mIU/mL.

### 2.7 Data analysis and statistics

RStudio (Version 1.2.5001), with R (version 3.6.1) was used for data analysis and figure generation. Additionally, the R add-on package “beeswarm” was utilised to visualise data as stripcharts with overlaying boxplots and to create non-overlaying data points. A second R add-on package “gplots” was used to generate specific colours for plots. Figures were exported from RStudio and then edited using Inkscape (Inkscape 1.2). Spearman’s rho coefficient was calculated to determine correlation between IGRA results and ACE2 binding inhibition using the “cor” function from R’s “stats” library. Mann-Whitney-U test and Wilcoxon test were used to determine difference of signal distributions between dialysed and non-dialysed groups for unpaired and paired samples, respectively using the “wilcox.test” function from R’s “stats” library. To assess differences in the study population, Pearson’s Chi-squared test with Yates’ continuity correction was used for categorical characteristics using the “chisq.test” function from R’s “stats” library and Mann-Whitney-U test as above was used for difference in age. The type of statistical analysis performed (when appropriate) is listed in the figure legends. Pre-processing of data such as matching sample metadata and collecting results from multiple assay platforms was performed in Excel 2016.

## 3. Results

### 3.1 Inferior humoral responses in haemodialysis patients after triple BNT162b2 vaccination

To characterise the vaccination response after the third BN162b2 vaccination in 50 patients on maintenance haemodialysis, we had followed immunoglobulin levels longitudinally after the second dose of BNT162b2 using MULTICOV-AB, a multiplex immunoassay containing antigens from the Spike protein of SARS-CoV-2 and selected variants of concern (9). As a novel control group, 33 samples from healthcare workers with triple BNT162b2 vaccination were used for comparison. Detailed information on the study populations can be found in Table 1, S1 and S2. Consistent with our previous reports (16, 17), IgG responses towards the original B.1 isolate in vaccinated dialysis patients were significantly reduced (p=4·68*10^−5^, Mann-Whitney-U test) when compared to the control group and declined after the second vaccination to comparable levels in both groups (p=7·33*10^−2^, Mann-Whitney-U test, Fig. 2a). A third BNT162b2 vaccination about six to eight months after the second increased the peak IgG RBD B.1 response in both groups but with higher variability in dialysis patients (p=4·02*10^−2^, Mann-Whitney-U test, Fig. 2a). As an additional control, quantitative S1 IgG titres were measured using a commercial assay (Fig. S2), which led to a very similar pattern of significantly diminished antibody responses in dialysis patients compared to non-dialysed individuals after the second BNT162b2 dose, declining titres and a robust peak response increase after the third vaccination.

**Fig. 2.**
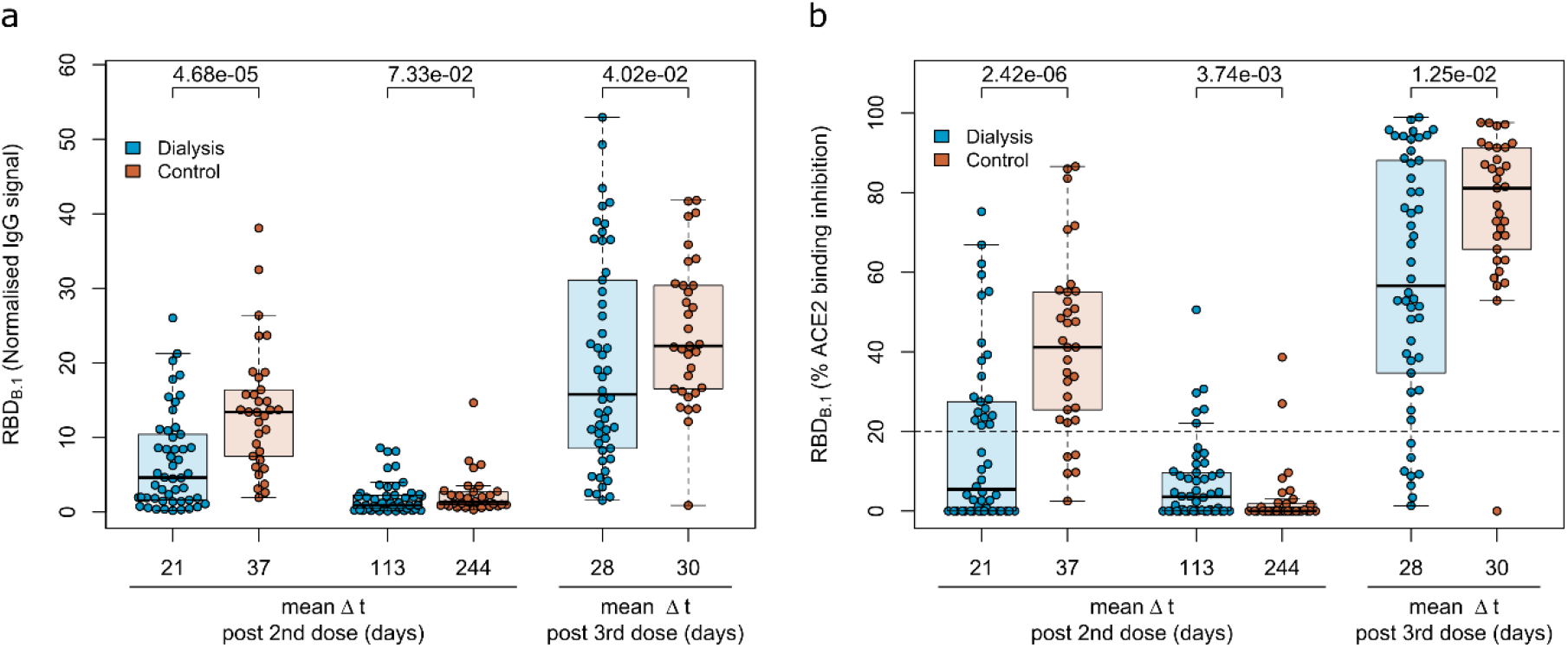
Humoral immune response haemodialysis patients after a triple vaccination with BNT162b2. IgG response (a) and ACE2 binding inhibition (b) towards the SARS-CoV-2 B.1 RBD isolate were measured in plasma from haemodialysis patients (blue circles, n=50) and controls (orange circles, n=33) using MULTICOV-AB (a) or an ACE2-RBD competition assay (b) after double or triple vaccination with BNT162b2 at the indicated time points. Data is displayed as normalised median fluorescence intensity (MFI) signal (a) for IgG binding or as % ACE2 binding inhibition where 100% indicates maximum inhibition and 0% no inhibition (b). Samples with an ACE2 binding inhibition of less than 20% (dashed line) are classified as non-responders. Boxes represent the median, 25th and 75th percentiles, whiskers show the largest and smallest non-outlier values. Outliers were determined by 1·5 times IQR. Mean sampling time in days after two-dose BNT162b2 vaccination as Δt is displayed on the x-axis. Statistical significance was calculated by two-sided Mann-Whitney-U test. P-values for relevant comparisons are given above the sample groups. Significance was defined as p<0·05. Response data from dialysed individuals from day 21 and day 113 after the second BNT162b2 dose were already published before as part of Strengert *et al*. (16) and Dulovic *et al*. (17).

For a functional characterisation of vaccine-induced antibodies towards the original B.1 RBD isolate, RBDCoV-ACE2, we used a multiplex competitive inhibition assay (23). ACE2 binding inhibition was significantly reduced in dialysed compared to non-dialysed individuals (p=2·42*10^−6^, Mann-Whitney-U test) after the second vaccination (Fig. 2b). Responses were comparably diminished in both groups four to eight months after the second vaccination, with only 12% and 6% of samples being above the 20% responder threshold in patients on haemodialysis and controls, respectively. However, comparable to IgG binding levels, the third BNT162b2 vaccination restored and even augmented ACE2 binding inhibition against the B.1 variant in both populations.

### 3.2 Strong immune responses after a fourth mRNA-1273 vaccination in haemodialysis patients

Next, we followed the anti-Spike RBD IgG levels in haemodialysis patients after the third vaccination over time and after a fourth vaccination with a full 100 µg dose of mRNA-1273, which was considered by German guidelines for immunocompromised individuals. As expected, IgG responses against the original B.1 isolate had again declined within approximately 4 months after the third vaccination (Fig. 3a, Fig. S2) as did the ACE2 binding inhibition activity as a surrogate for virus neutralisation (Fig. 3b). While the decline was not as severe as after the second BNT162b2 dose with now 64% of samples remaining above the 20% ACE2 binding inhibition threshold, only the fourth vaccination with mRNA-1273 markedly raised both anti-Spike RBD IgG levels (Fig. 3a, Fig. S2, Table S3 for a complete statistical evaluation) and ACE2 binding inhibition (Fig. 3b) towards the B.1 isolate above levels seen at peak response after the second and third dose of BNT162b2. 96% of samples from individuals on haemodialysis were now classified as above the 20% ACE2 responder threshold. Further, we also analysed the longitudinal development of ACE2 binding inhibition towards the dominantly circulating SARS-CoV-2 of 2021 (Delta) and 2022 (Omicron) (Fig 4c, d). ACE2 binding inhibition towards the Delta Variant was slightly reduced over time to levels observed with the B.1 isolate. Overall, the third dose resulted in a clear increase in Delta ACE2 responder rates from 24% after two-dose BNT162b2 scheme to 64%, which was further increased to 94% after the subsequent dose of mRNA-1273 (Fig. 4c). Importantly, neutralisation against the Omicron BA.1 variant, which was largely absent after the second vaccination and only transiently above threshold in 38% of dialysis patients after the third vaccination, reached high levels of ACE2 binding inhibition with an 80% responder rate at peak response after the fourth vaccination with mRNA-1273, which coincided with Omicron being the dominant SARS-CoV-2 variant circulating in Germany (Fig. 4d).

**Fig. 3.**
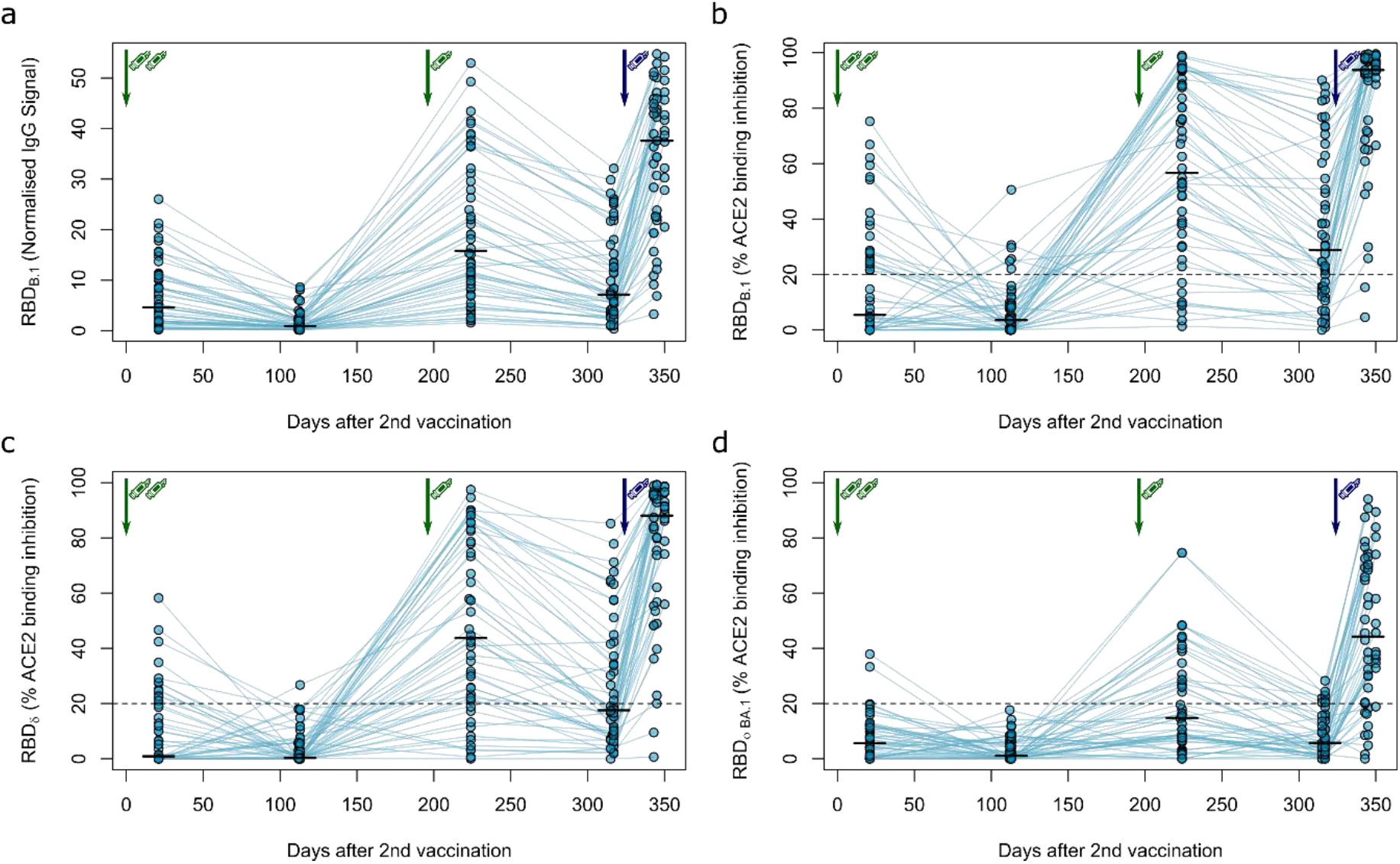
Longitudinal humoral immune response in haemodialysis patients after a triple vaccination with BNT162b2 and a fourth full-dose of mRNA-1273. IgG response (a) and ACE2 binding inhibition (b, c, d) towards the SARS-CoV-2 RBD of B.1 (a, b), δ (c) and Ο BA.1 (d) isolates were measured in plasma from haemodialysis patients (n=50) using MULTICOV-AB (a) or an ACE2-RBD competition assay (b, c, d) after immunisation with a triple dose of BNT162b2 (green syringe) and a fourth full-dose of mRNA-1273 (blue syringe). Data is displayed as normalised median fluorescence intensity (MFI) signal for IgG binding (a) or as % ACE2 binding inhibition where 100% indicates maximum inhibition and 0% no inhibition (b, c, d). Samples with an ACE2 binding inhibition of less than 20% (dashed line) are classified as non-responders (b, c, d). Interconnecting lines represent samples from the same individual. Sampling time points in days after the standard complete two-dose BNT162b2 vaccination is stated below the graph. Statistical significance was calculated by two-sided paired Wilcoxon rank test. Significance was defined as p<0·05. All p-values for relevant comparisons are listed in Table S3.

**Fig. 4.**
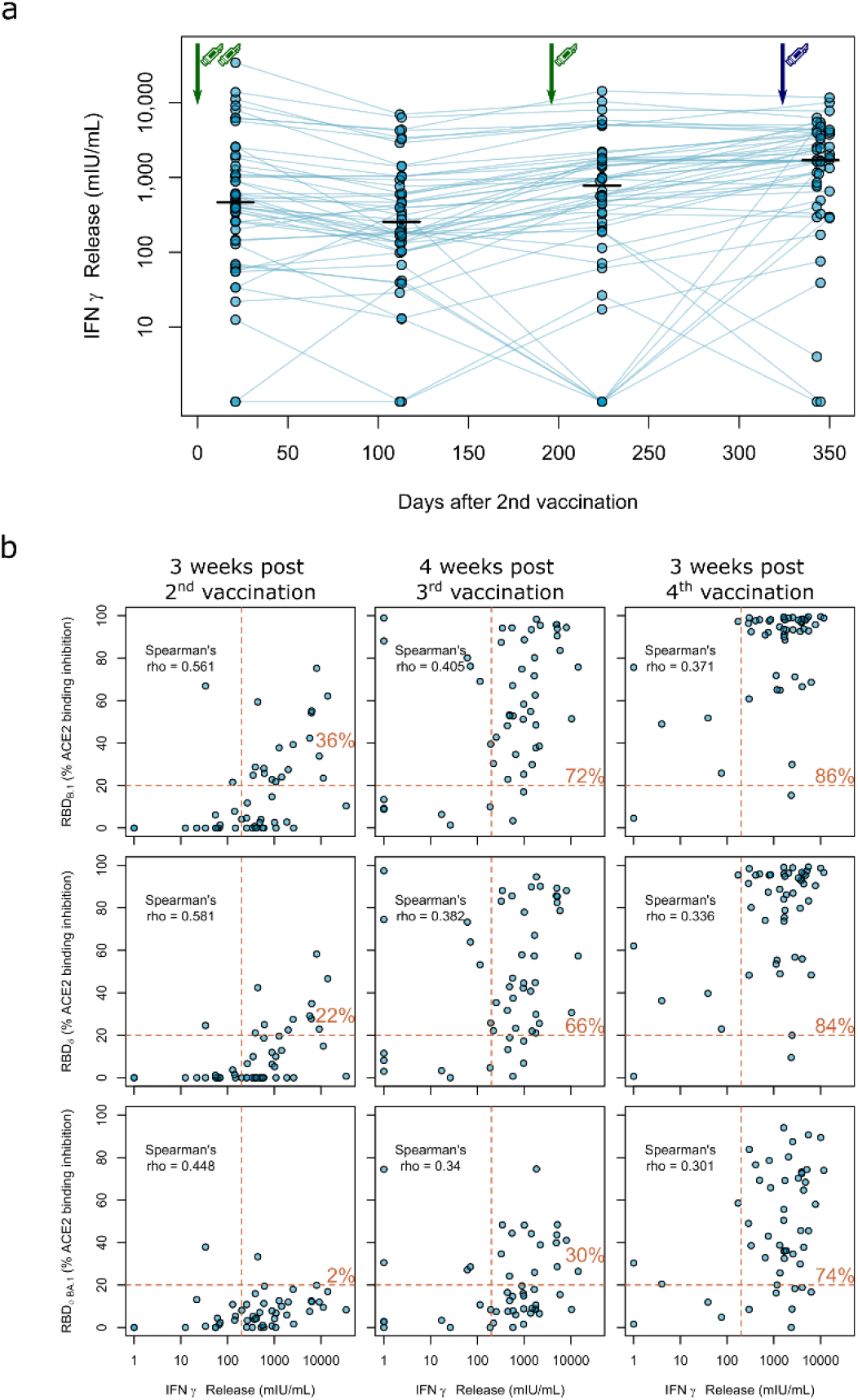
Impact of triple vaccination with BNT162b2 and a fourth full-dose of mRNA-1273 on cellular immune response in haemodialysis patients. (a) Whole blood from longitudinally-sampled vaccinated haemodialysis patients (n=50) was *ex vivo* stimulated using a SARS-CoV-2 Spike S1-specific peptide pool. Supernatant fractions were then analysed by interferon γ release assay (IGRA). Interconnecting lines represent samples from the same individual. Sampling time points in days after two-dose BNT162b2 vaccination is displayed on the x-axis. Statistical significance was calculated by two-sided paired Wilcoxon rank test. Significance was defined as p<0·05. All p-values for relevant comparisons are listed in Table S3. Response data from dialysed individuals from day 21 after the second BNT162b2 dose were already published before as part of Strengert *et al*. (16) and Dulovic *et al*. (17). (b) T-cell responses assessed by IGRA and B-cell responses assessed by ACE2-RBD competition assay towards the RBD of B.1, Delta, Omicron BA.1 isolates were plotted for correlation analysis (b). Correlation was calculated using Spearman’s coefficient rho. Dashed lines indicated the respective responder thresholds for IGRA (IFNγ release >200 mIU/mL) and RBD-ACE2 binding assay (20%). Responder rates (%) for both cellular and humoral response are shown in the upper right quadrant.

We also analysed IgG binding longitudinally after a triple dose of BNT162b2 towards the RBD of B.1, Delta and Omicron BA.1 VoC in saliva of haemodialysis patients to determine protection levels at the primary side of SARS-CoV-2 replication. Although anti-RBD specific IgG was readily detectable both in the peak and plateau response phase following the complete two-dose and the third booster dose of BNT162b2, IgG binding towards the Delta and Omicron BA.1 RBD was significantly reduced compared to the B.1 RBD across all time points (Fig. S3). Interestingly, saliva responses across vaccinated individuals were much more widespread in saliva than in plasma.

As clinical studies suggested that both cellular and humoral response can confer protection from COVID-19 (25), we also assessed vaccination-induced T-cell responses by IFNγ release assay longitudinally. Overall, these responses were more stable over time (Fig. 4a). After two BNT162b2 vaccinations, IFNγ release after *in vitro* re-stimulation was readily detectable in haemodialysis patients, but declined slightly thereafter. The third BNT162b2 vaccination increased cellular responses to levels comparable to after the second vaccination. Similar to the humoral responses, the fourth vaccination with mRNA-1273 further increased IFNγ release after Spike S1 peptide restimulation of T-cells (Fig. 4a, Table S3 for a complete statistical evaluation).

Finally, we correlated B- and T-cell responses after each vaccination within our longitudinal cohort of haemodialysis patients. We overall observed moderate correlation between peak T-cell responses (measured by IGRA) and B-cell responses [determined by % ACE2 binding inhibition of the B.1 variant (Spearman’s rho=0·561, Fig. 4b, upper panel)], which did not increase after the third (Spearman’s rho=0·405) and fourth (Spearman’s rho=0·371) vaccination. We further described responder rates for T- and B-cell response by a combined cut-off as displayed in Fig. 4b. Notably, responder rates among haemodialysis patients strongly increased to 72% after the triple BNT162b and further to 86% after the fourth full-dose mRNA-1273 dose. Importantly, whilst we observed a similar trend for the correlation coefficient between Delta and Omicron BA.1 % ACE2 binding inhibition and T-cell responses (Fig. 4b; middle and lower panel, Table S3 for a complete statistical evaluation), dual cellular and humoral responders levels equally strongly increased for both VoCs after the third and fourth vaccination to a final 84% and 74%, respectively.

## 4. Discussion

Although overall case mortality rates for SARS-CoV-2 have significantly decreased since the initial wave of the pandemic, maintaining high levels of vaccine-induced protection is of paramount importance for at-risk individuals for severe COVID-19 such as haemodialysis patients. Ensuring that these and other similarly vulnerable individuals are sufficiently protected remains challenging, with high case numbers throughout 2022 as a result of successive occurrence of Omicron subvariants. Despite clear recommendations on the need for a fourth dose, worryingly this fourth dose uptake among haemodialysis patients has decreased compared to the first three doses, with disparities among demographic groups remaining in place (26). At present, recommendations by the German Standing Committee on Vaccination (STIKO) clearly endorse a fourth SARS-CoV-2 vaccine dose including a full dose of mRNA-1273 for immunocompromised individuals (5), which contrasts WHO guidelines recommending 50 µg mRNA-1273 for booster vaccinations (27).

Several studies report of superior immunity after initial mRNA-1273 prime/boost vaccination when compared to BNT162b2 in haemodialyis patients (28, 29) or in the general population (30-32) and further improved humoral responses after triple vaccination in dialysis patients (33-37). Third dose vaccination with mRNA-1273 or BNT162b2 vaccines provided comparable protection against symptomatic SARS-CoV-2 infection in the general population although differences between both vaccines were observed after the second dose (38). Finally, Caillard et *al*. found that a four-dose mRNA-1273 compared to a four-dose BNT162b2 results in increased levels of binding antibodies in kidney transplant recipients (39).

We can only speculate about the effects of mixing mRNA-based vaccines. Janssen *et al*. compared heterologous and homologous mRNA-1273 and BNT162b2 vaccination after the respective first vaccination in a randomized trial (40). They found the geometric mean titers of anti-spike IgG antibodies for each heterologous regimen to be higher relative to the corresponding homologous regimen. This is consistent with data from Israel (41) and the COV-BOOST study (42), in which even half-dose mRNA-1273 as fourth dose after triple BNT162b2 vaccination appeared to have higher immunogenicity than full-dose BNT162b2. The authors suggested that this result might be due to a heterologous schedule effect or the vaccine dose. Interestingly, differences between both mRNA vaccines could be more complex, since mRNA-1273 is reported to induce higher concentrations of RBD- and N-terminal domain-specific IgA and more antibodies eliciting neutrophil phagocytosis and natural killer cell activation as compared to BNT162b2 (43).

Our study is, to our knowledge, the only study examining the longitudinal humoral and cellular immune response towards the most recent SARS-CoV-2 isolates in haemodiaylsis patients after administration of consistent vaccination regimens starting with a triple dose of BNT162b2 followed by a fourth full-dose of mRNA-1273. Whilst other studies principally support the beneficial impact of a fourth vaccination dose on both antibody titers and neutralizing potency towards SARS-CoV-2 B.1 and VoC isolates, often various vaccination regimens including heterologous vector-based/mRNA regimens were pooled in cohorts (14) or vaccine dosages not provided (13).

Our data provide solid evidence that the triple vaccination resulted in mean antibody concentration and neutralizing activity above levels to after the second vaccination. Interestingly, we identified significant further increases in both humoral and cellular response rates following the fourth dose, compared to the second and third. The increase in response rate from 30% to 74% from third to fourth dose for Omicron is particularly important considering it comprises almost all currently circulating variants of SARS-CoV-2. We consider this as a valid argument for a fourth vaccination in at-risk patients, especially, since T-cell immunity elicited by current vaccines is also effective against VoC including Omicron (44-46). The large range in both humoral and cellular responses illustrates however, the variable nature of SARS-CoV-2 vaccination responses in dialysis patients and maybe of relevance for identifying individuals with inferior responses in need for further doses.

Our study has several limitations. The number of participants within our cohort was limited, with only 50 patients on haemodialysis and a further 33 control participants, although our sample size is larger than similar studies examining the effect of the fourth dose within haemodialysis patients (15). The use of longitudinal cohort also allows us to directly identify the responses and their decline following each individual dose. Unfortunately, we were unable to obtain samples post-fourth dose for our control population, since additional booster vaccinations are not generally recommended and a full dose mRNA-1273 vaccination would be the unlikely regimen for the healthy controls. Although our control group was well-matched for sample collection at peak antibody levels after the second and third vaccination, they were not optimally matched for age and gender. Finally, it would have been interesting to directly compare homologous fourth BNT162b2 dose to mRNA-1273 in haemodyalysis patients and to assess the reactogenicity, but this would have required a prospective study design for an interventional study.

Overall, a fourth full-dose of the mRNA-1273 vaccine elicits improved cellular and humoral responses compared to the triple BNT162b2 vaccination and appears to be an advisable strategy for immunocompromised patients, such as haemodialysis patients. Nevertheless, the decline after fourth vaccination and the effectivity against emerging SARS-CoV-2 variants will have to be monitored to assess the immune response duration and requirement for further booster vaccinations.

## Contributors

GMNB, NSM, AD and MS conceived the study. MB, AD, MS, AD-J, GMNB, AC, NSM, DJ and MVS designed the experiments. NSM, MS, GMNB, AD-J, and GK procured funding. GMR, JG, JJ, DJ and MVS performed experiments. KL, AB, EW, GL, AC, and GMNB collected samples or organised their collection. PDK, BT and UR produced and designed recombinant assay proteins. MB, KL, AD, MS, GMR, MVS and AC performed data collection and analysis. MB generated the figures. MB, MS, AD and GMNB verified the underlying data. GMNB and MS wrote the first draft of the manuscript with input from MB, AC, KL and AD. All authors critically reviewed and approved the final manuscript.

## Supporting information

Supplementary Material Becker et al. 2022

## Data Availability

Data relating to the findings of this study are available from the corresponding authors upon request.

## Acknowledgments

We sincerely thank all patients for their continued contribution and willingness to participate in this study. We also thank all clinical staff at the Eickenhof Dialysis Centre for their efforts to make this study possible.

