## Supplementary Material Becker et al. 2022 for "Longitudinal cellular and humoral immune responses after triple BNT162b2 and fourth full-dose mRNA-1273 vaccination in haemodialysis patients"

Author Affiliations

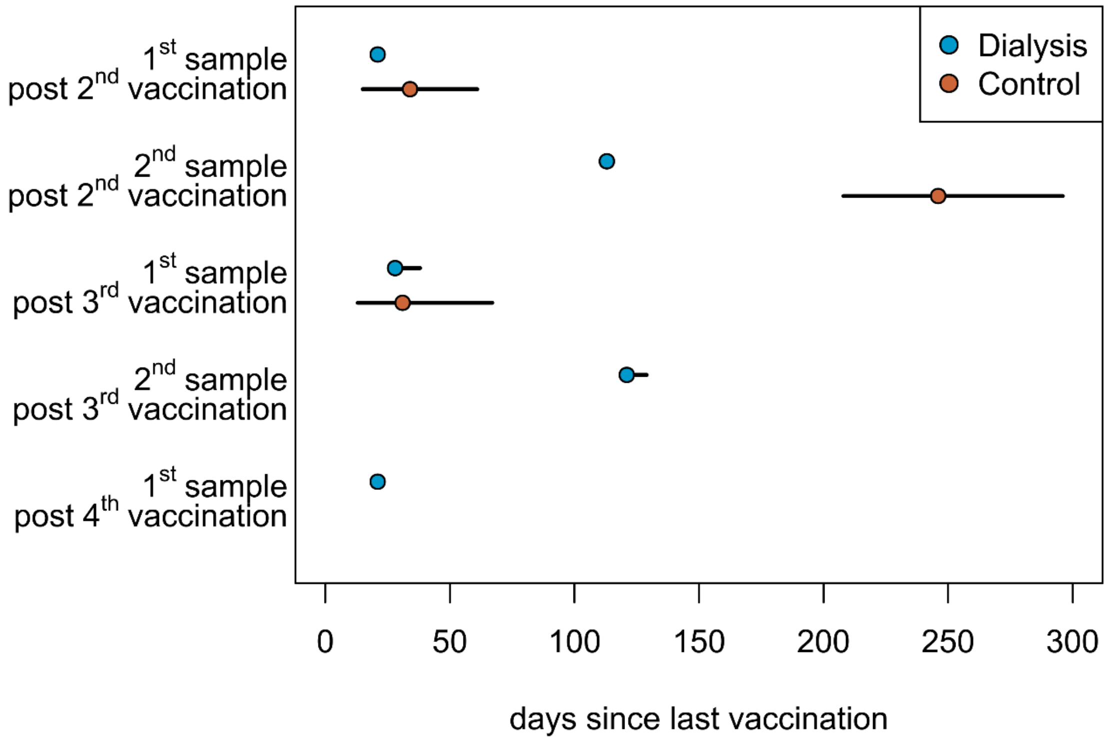


**Fig. S1. Sampling time points after COVID-19 vaccination in the study population.**
Graphic display of median and absolute range of sampling times after the indicated vaccination of haemodialysis patients (n=50, blue circles) and healthcare workers (n=33, orange circles), who served as controls. Controls were triple-vaccinated with BNT162b2 (vaccination 1-3) whereas haemodialysed individuals received an additional full-dose of mRNA-1273 (vaccination 4).

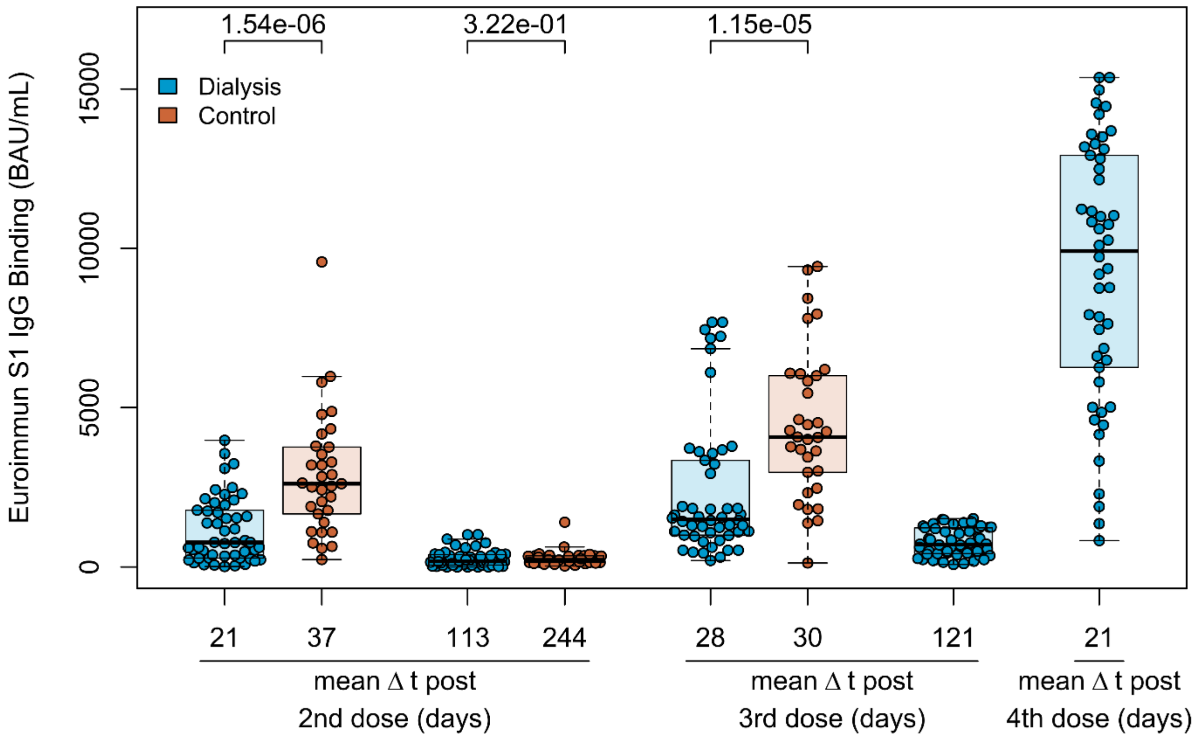


**Fig. S2. Development of quantitative plasma IgG titres after COVID-19 vaccination.** Spike subdomain 1 (S1)-plasma IgG from haemodialysis patients (blue circles, n=50) and controls (orange circles, n=33) were analysed using the QuantiVac-ELISA from Euroimmun (BAU/mL) after a triple vaccination with BNT162b2. For haemodialysed individuals, S1 IgG titres are additionally shown after a fourth 100 µg (full) dose of mRNA-1273. Mean sampling time in days after the respective vaccination is displayed as Δt on the x-axis. Boxes represent the median, 25th and 75th percentiles, whiskers show the largest and smallest non-outlier values. Outliers were determined by 1·5 times IQR. Statistical significance was calculated by two-sided Mann-Whitney-U test. P-values for relevant comparisons are given above the sample groups. Significance was defined as p<0·05. Response data from dialysed individuals from day 21 and day 113 after the second BNT162b2 dose were already published before as part of Strengert *et al.* (1) and Dulovic *et al.* (2).

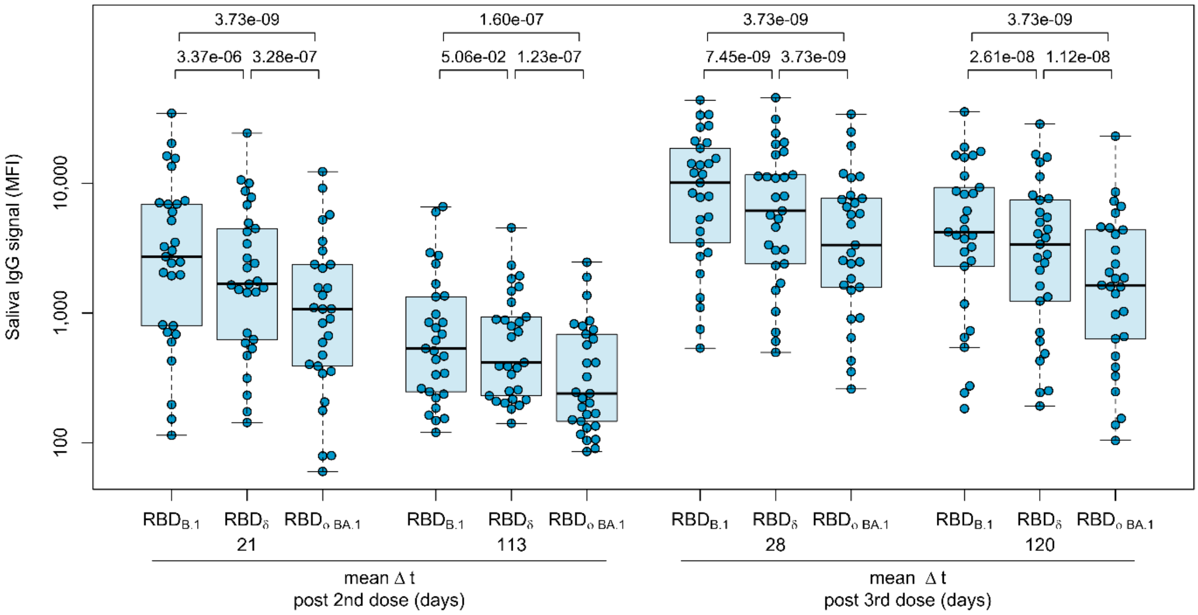


**Fig. S3. Mucosal immune response in haemodialysis patients after triple vaccination with BNT162b2.** IgG response in saliva of haemodialysis patients (n=29) towards the SARS-CoV-2 RBD of B.1, Delta and Omicron BA.1 isolates were measured using MULTICOV-AB. Data is displayed as median fluorescence intensity (MFI) signal for IgG binding. Sampling time points in days after a completed standard two-dose BNT162b2 vaccination is stated on the x-axis. Statistical significance was calculated by two-sided paired Wilcoxon rank test. Significance was defined as p<0·05. P-values for relevant comparisons are given above the sample groups. Response data from dialysed individuals from day 21 and day 113 after the second BNT162b2 dose were already published before as part of Strengert *et al.* (1) and Dulovic *et al.* (2).

**Table S1.** Indication for haemodialysis in the study population.

| Characteristics | Haemodialysis group (n=50) |
|---|---|
| Diagnosis (n, %) | |
| Autosomal dominant polycystic kidney disease | 7 (14·0) |
| Chronic glomerulonephritis | 3 (6·0) |
| Diabetic nephropathy | 8 (16·0) |
| Focal segmental glomerulosclerosis | 3 (6·0) |
| IgA nephropathy | 7 (14·0) |
| Interstitial nephritis | 4 (8·0) |
| Nephrosclerosis | 11 (22·0) |
| Acute toxic tubular epithelial damage syndrome | 1 (2·0) |
| Anti-Neutrophilic Cytoplasmic Autoantibody (ANCA)-associated vasculitis | 1 (2·0) |
| Medullary cystic kidney disease | 1 (2·0) |
| Membranous glomerulonephritis | 1 (2·0) |
| Kidney dysplasia | 1 (2·0) |
| Obstructive nephropathy | 1 (2·0) |
| Cystic kidney disease | 1 (2·0) |
| Cyclosporin intoxication | 1 (2·0) |

**Table S2.** Medication of study participants. NA - Information not available.

| Medication (n, %) | Haemodialysis group (n=50) | Non-dialysis control group (n=33) |
|---|---|---|
| Angiotensin-converting enzyme inhibitor | 15 (30·0) | 2 (6·1) |
| Statins | 28 (56·0) | 0 (0·0) |
| Angiotensin II Receptor Blocker | 16 (32·0) | 1 (3·0) |
| Vitamin D Supplements | 49 (98·0) | NA |
| L-Thyroxine | 0 (0·0) | 3 (9·1) |
| $Ca^{2+}$ channel antagonist | 0 (0·0) | 2 (6·1) |
| 5-aminosalicylic acid | 0 (0·0) | 1 (3·0) |
| DPP4 inhibitor + metformin | 0 (0·0) | 1 (3·0) |
| Factor Xa inhibitor | 0 (0·0) | 1 (3·0) |
| Immunosuppressants (dosing range per day) | | |
| Prednisolone (2 mg; every second day ) | 1 (2·0) | 0 (0·0) |
| Prednisolone (5 mg)* | 2 (4·0) | 0 (0·0) |
| Prednisolone (7·5 mg) | 1 (2·0) | 0 (0·0) |
| Prednisolone (5 mg), Tacrolimus (1-2 mg) | 2 (4·0) | 0 (0·0) |
| Prednisolone (5 mg), Tacrolimus (12 mg), Mycophenolatmofetil (500 mg)** | 1 (2·0) | 0 (0·0) |
| Hydrocortisone (20 mg) | 1 (2·0) | 0 (0·0) |
| 5-Fluorouracil*** | 1 (2·0) | 0 (0·0) |

* One patient discontinued prednisolone 128 days after the second and 68 days before the third BNT162b2 dose
** Mycophenolatmofetil discontinued 166 days after the second and 30 days before the third BNT162b2 dose
*** First treatment cycle started 63 days before the fourth vaccination with mRNA-1273

**Table S3.** Statistical comparison of longitudinal cellular and humoral vaccination responses in haemodialysis individuals.

| Assay | Variant | Figure | Sample 1 vs 2 | Sample 1 vs 3 | Sample 3 vs 4 | Sample 3 vs 5 |
|---|---|---|---|---|---|---|
| MULTICOV-AB | B.1 | 3a | $1·51*10^{-9}$ | $1·61*10^{-9}$ | $2·44*10^{-9}$ | $1·05*10^{-9}$ |
| RBDCoV-ACE2 | B.1 | 3b | 0·005 | $7·79*10^{-10}$ | $4·38*10^{-9}$ | $7·79*10^{-10}$ |
| RBDCoV-ACE2 | Delta | 3c | 0·013 | $1·15*10^{-9}$ | $1·20*10^{-8}$ | $7·79*10^{-10}$ |
| RBDCoV-ACE2 | Omicron BA.1 | 3d | 0·002 | $3·52*10^{-7}$ | $8·48*10^{-7}$ | $1·15*10^{-9}$ |
| IGRA | B.1 | 4a | $1·38*10^{-5}$ | 0·564 | NA | 0·0005 |
